# Risk of Suicidal Ideation and Behaviors, Depression, and Anxiety with GLP-1 Receptor Agonist Use in Children and Adolescents: A Target Trial Emulation Study

**DOI:** 10.1101/2025.11.11.25339931

**Authors:** Pareeta Kotecha, Yao An Lee, Angelina V Bernier, Sarah C Westen, Steven M Smith, Pengyue Zhang, Tamara S. Hannon, Jiang Bian, Jingchuan Guo

## Abstract

**Importance:** Rising rates of obesity and youth-onset type 2 diabetes (YT2D) in children and adolescents have increased GLP-1 receptor agonist (GLP-1 RA) use, raising concerns about potential effects on suicidal ideation/behaviors (SI/SB), depression, and anxiety.

**Objective:** To assess associations between GLP-1 RA use for obesity or YT2D and risk of SI/SB, depression, and anxiety in children and adolescents.

**Setting:** OneFlorida+ EHR from January 1, 2020, to January 31, 2024.

**Design:** Retrospective cohort study using prevalent-new user design, target trial emulation framework, and sIPTW for confounding control.

**Participants:** Children and adolescents with obesity or YT2D who were ≥6 and <18 years were included with no history of the respective outcomes.

**Exposure:** New GLP-1 RA users vs. Prevalent metformin users.

**Outcomes:** SI/SB, anxiety, and depression. Weighted Cox proportional hazards models were used to assess the risk of outcomes. Risk differences (RD) and hazard ratios (HR) are presented with 95% CI. RDs are per 1,000 person-years.

**Results:** The study included 2,010, 1,774, and 1,764 patients for SI/SB, depression, and anxiety, respectively. The mean age was ∼14.2 years, ∼61% were female, with up to 4 years of follow-up, across cohorts. Compared to prevalent metformin users, GLP-1 RA users had lower incidence and risk of SI/SB (RD: -10.45, -14.54 to -6.36; HR: 0.11, 0.02 to 0.86) and depression (RD: -25.64, -34.90 to -16.39; HR: 0.37, 0.17 to 0.78). There was no difference in anxiety risk between the two groups (RD: 5.95, -7.10 to 19.01; HR: 1.13, 0.69 to 1.84).

**Conclusions:** GLP-1 RA use may reduce the risks of SI/SB and depression among children and adolescents with obesity or YT2D; no association was found with anxiety. Long-term surveillance is needed.

**KEY POINTS:** *Question:* Are GLP-1 receptor agonists (GLP-1 RAs) for the indication of obesity or youth-onset type 2 diabetes (YT2D) associated with the risk of mental health outcomes in children and adolescents?

*Findings:* In this retrospective cohort study, including 2,116 children and adolescents with obesity or T2D, GLP-1 RA use was associated with a significantly lower risk of suicidal ideation and behaviors and depression when compared to prevalent metformin users. There was no significant difference in the risk of anxiety between the two groups.

*Meaning:* The results of this study suggest that children and adolescents using GLP-1 RA for obesity or YT2D were at lower risk of suicidal ideation and behaviors, and depression compared to prevalent metformin users. The risk of anxiety remained non-significant, but the higher trend warrants future surveillance by pediatricians, psychologists, and pharmacoepidemiologists.

## INTRODUCTION

Childhood overweight (BMI ≥ 85th percentile to <95th percentile) and obesity (BMI ≥ 95th percentile) are persistent, chronic health conditions that have reached epidemic levels, affecting 16.1% of children aged 2 to 9 years and 19.3% of those aged 10 to 19 years.^1^ This rise in cardiometabolic disease in youth, including youth-onset type 2 diabetes (YT2D) has paralleled the increases in pediatric obesity, with projections indicating a continued rise in the incidence rates and prevalence of YT2D.^2,3,4^ In addition to YT2D, children and adolescents with obesity are at elevated risk for comorbidities such as liver disease, hypertension, dyslipidemia, obstructive sleep apnea, cancer, reproductive health issues, and earlier onset of cardiovascular disease, potentially leading to a longer duration of chronic disease burden across their lifespan.^1,5^ Given the public health burden, early intervention in children and adolescents with obesity and YT2D is crucial.

Lifestyle interventions remain the primary approach to treat pediatric and adolescent obesity; however, pharmacotherapy is recommended to manage both obesity and YT2D.^1^ Despite this need, treatment options have remained limited to metformin and insulin for over three decades before glucagon-like peptide-1 receptor agonists (GLP-1 RAs) were approved for YT2D and/or obesity treatment in children and adolescents.^1,6,7^ This therapeutic gap has contributed to the increased use of newer pharmacologic agents, particularly GLP-1 RAs, between 2020 and 2023.^8^ Currently, the U.S. Food and Drug Administration (FDA) and the European Medicines Agency (EMA) have approved liraglutide and semaglutide for adolescents aged ≥12 years with obesity, while liraglutide, dulaglutide, and exenatide are approved for YT2D.^1,9^ Tirzepatide, a dual GLP-1/GIP receptor agonist, is under active investigation for this population.^10^

The psychological effects of GLP-1 RAs have recently become a topic of interest and concern. Regulatory bodies, including the FDA and the EMA, are currently investigating potential links between GLP-1 RA use and suicidal thoughts or behaviors.^11,12^ However, studies have largely remained limited to adults, with little focus on children and adolescents, and the results from studies in adults remain mixed.^13,14,15^ It is noteworthy that the suicidal ideation and behavior rates among children and adolescents have nearly tripled from 2007 to 2017, in the U.S, with 17% reporting suicidal ideation and 7-8% reporting a suicide attempt each year.^16^ Although the role of GLP-1 RAs in depression and anxiety has not been studied in clinical trials or assessed through real-world evidence in this population, there is a critical need to examine the potential neuropsychiatric effects of GLP-1 receptor agonist use in children and adolescents. This need is further underscored by the fact that an estimated 4% of U.S. children and adolescents have been diagnosed with depression, and 10% with anxiety disorders.^17,18^ Additionally, more than one in three children and adolescents with a current mental health condition had two or more co-occurring conditions, highlighting the complexity and potential vulnerability of this population.^18^

Given the critical need to understand the impact of GLP-1RAs on mental health outcomes in youth, this study aimed to assess the association between GLP-1 RA use for the indication of obesity or YT2D in children and adolescents and three psychological outcomes, including suicidal ideation and behaviors, depression, and anxiety. Our study sought to address the critical knowledge gaps regarding the real-world impact of GLP-1 RAs for obesity or YT2D on mental health outcomes in children and adolescents.

## METHODS

### Study Design, Data Source, and Study Population

This retrospective cohort study was conducted using OneFlorida+ Clinical Research Network electronic health record (EHR) data spanning from January 1, 2020, to January 31, 2024. OneFlorida+ is a large-scale, population-based research repository containing longitudinal EHRs for over 21 million individuals across Florida, Georgia, and Alabama. The database includes comprehensive patient-level information, including diagnoses, prescriptions, fills, procedures, vital signs, laboratory values, and death data, with linkage to the National Death Index. OneFlorida+ adheres to the PCORnet Common Data Model and is updated quarterly, ensuring interoperability and consistency for multisite research.^19^

Our study followed a target trial emulation framework and used a prevalent new-user design to evaluate the association of GLP-1 RAs with psychological outcomes in children and adolescents. We included children and adolescents with obesity or YT2D who were aged ≥6 and <18 years, and had ≥1 healthcare encounter in the year prior to cohort entry. Patients with YT2D were identified using ≥1 International Classification of Diseases (ICD) code for T2D, and patients with obesity were identified using at least 1 ICD code for obesity or a BMI percentile ≥95% (**eTable 1 in Supplement 1**).

Patients were excluded if they had a diagnosis of pregnancy, gestational diabetes, end-stage renal disease (ESRD), medullary thyroid carcinoma, multiple endocrine neoplasia type 2, active malignant neoplasm, or no visits after the prescription of the study exposures. Patients with the outcome of interest present at baseline were excluded from analyses of that specific outcome to ensure incident case identification.

### Exposure Definition

Patients were categorized as new GLP-1 RA users or prevalent metformin users identified by RxNorm and National Drug Codes. Prevalent metformin users were chosen as a comparator group based on the literature suggesting limited treatment options and clinical expertise.^6,7^ The pharmacological treatment options for children and adolescents with YT2D and obesity have largely remained limited over the years, leading to the use of metformin for YT2D as per the guidelines.^9^ A significant off-label use of metformin has been observed in this population with obesity and overweight due to its safety profile and low cost.^6^

GLP-1 RA users were defined as children or adolescents who initiated GLP-1 RA therapy on or after January 1, 2020, i.e., the year of the first GLP-1 RA approval for pediatric populations in the U.S. The index date, i.e., follow-up start date, was defined as the first date of GLP-1 RA prescription. The comparator group consisted of children and adolescents who did not initiate GLP-1 RAs at any time during the study period, but who were prevalent metformin users. To qualify, individuals had to have at least one metformin prescription during the baseline period, and the date of the subsequent metformin prescription during the eligible study period, i.e., during 2020-2024, was designated as the index date.

### Outcomes

Three co-primary outcomes were evaluated. These included incident (1) suicidal ideation or behaviors, (2) depression, and (3) anxiety. Each outcome was identified using validated algorithms with ICD-10 diagnosis codes, and the date of first diagnosis was captured as the outcome date.^20,21,22,23,24,25^ The ICD diagnosis codes used for the outcomes are detailed in **eTable 1 in Supplement 1**.

Suicidal ideation and behaviors encompassed suicidal ideation, suicidal behaviors, including intentional self-harm and self-injurious behaviors without suicidal intent. These were grouped per the U.S FDA guidance for industry for post-marketing surveillance of suicidal ideation and behaviors.^20^

Depression and anxiety were identified using ICD codes as per the American Psychiatric Association’s Diagnostic and Statistical Manual of Mental Disorders, 5th edition (DSM-5).^24,25^

### Covariates

Baseline comorbidities were assessed up to three years before the index date, and sociodemographics, clinical observations, and co-medications were assessed up to 1 year before or on the index date. Sociodemographic covariates included age, sex, race/ethnicity, and insurance type. Clinical variables included recent body mass index (BMI), BMI standard deviation score (BMI-SDS/BMI z-score), and BMI percentiles. BMI-z scores and BMI percentiles were calculated using the Centers for Disease Control and Prevention’s (CDC) 2022 extended growth charts for those with obesity and the 2000 Growth Charts for those without obesity based on BMI, height, weight, and other metrics as required by the CDC.^26^ Additionally, we included recent lab results for HbA1c as binary variables for HbA1c <6.5%, 6.5-8.9%, and ≥9%. Comorbidities assessed included a wide range of metabolic, psychiatric, developmental, and chronic conditions, specific to children and adolescents, and were identified using ICD codes.

Overweight was identified using ICD codes or BMI percentile ≥85% and <95%. We also calculated the pediatric comorbidity index score, calculated using a validated macro by Sun et al, operationalized as a score of <4 or ≥4.^27^ Medication history in the 12 months prior to the index date was captured for antidiabetic, anti-obesity, antihypertensive, lipid-lowering, cardiovascular, psychological, and other commonly prescribed medication classes identified using National Drug Codes and RxNorm codes. Other covariates included healthcare utilizations, such as primary care physician, psychiatrist, psychologist, counseling, pediatrician, and dietary and exercise intervention visits with the nutritionist.

### Statistical Analysis

We applied stabilized inverse probability of treatment weighting (sIPTW) to control for potential confounding due to non-randomized treatment allocation. This method generated a pseudo-population in which the distribution of observed baseline characteristics was independent of treatment assignment.^28^ The sIPTW weights were derived from propensity scores (PS), estimated through a multivariable logistic regression model predicting the likelihood of initiating GLP-1RA therapy, using the covariates listed in **Table 1**.^28^

**Table 1:** Baseline Characteristics of GLP-1 RA Users vs. Prevalent Metformin Users Within the 3 Study Cohorts.

| Variables | Suicidal Ideation and Behaviors |  |  |  | Depression |  |  |  | Anxiety |  |  |  |
| --- | --- | --- | --- | --- | --- | --- | --- | --- | --- | --- | --- | --- |
|  | Treatment (Total N = 2010) |  | SMD |  | Treatment (Total N= 1774) |  | SMD |  | Treatment (Total N= 1764) |  | SMD |  |
|  | Prevalent Metformin Users | GLP-1 RA Users | Pre-IPTW | Post -IPTW | Prevalent Metformin Users | GLP-1 RA Users | Pre-IPTW | Post -IPTW | Prevalent Metformin Users | GLP-1 RA Users | Pre-IPTW | Post-IPTW |
|  | (N=1461) | (N=549) |  |  | (N=1291) | (N=483) |  |  | (N=1281) | (N=483) |  |  |
| Age in years | 13.7±2.5 | 14.7±2.1 | 0.43 | -0.04 | 13.6±2.5 | 14.6±2.2 | 0.41 | -0.04 | 13.7±2.5 | 14.6±2.2 | 0.4 | -0.03 |
| Female | 901 (61.7) | 334 (60.8) | 0.02 | 0 | 800 (62) | 290 (60) | 0.04 | -0.01 | 797 (62.2) | 290 (60.0) | 0.04 | -0.03 |
| Race/Ethnicity |  |  |  |  |  |  |  |  |  |  |  |  |
| Non-Hispanic White | 346 (23.7) | 114 (20.8) | -0.07 | -0.05 | 308 (23.9) | 98 (19.6) | -0.1 | -0.08 | 298 (23.3) | 83 (17.2) | -0.15 | -0.04 |
| Non-Hispanic Black | 385 (26.4) | 156 (28.4) | 0.05 | 0.13 | 346 (26.8) | 142 (28.5) | 0.04 | 0.12 | 340 (26.5) | 146 (30.2) | 0.08 | 0.13 |
| Hispanic | 591 (40.5) | 220 (40.1) | -0.01 | -0.09 | 513 (39.7) | 204 (40.9) | 0.02 | -0.08 | 524 (40.9) | 203 (42.0) | 0.02 | -0.11 |
| Other Race | 50 (3.4) | 16 (2.9) | -0.03 | 0.04 | 47 (3.6) | 14 (2.9) | -0.03 | 0.08 | 44 (3.4) | 14 (2.9) | -0.03 | 0.01 |
| Unknown Race | 89 (6.1) | 43 (7.8) | 0.07 | 0 | 77 (6.0) | 39 (8.1) | 0.08 | 0.01 | 75 (5.9) | 37 (7.7) | 0.07 | 0.03 |
| Insurance Type |  |  |  |  |  |  |  |  |  |  |  |  |
| Medicaid | 718 (49.1) | 261 (47.5) | -0.03 | 0.05 | 626 (48.5) | 235 (48.6) | -0.01 | 0.06 | 632 (49.3) | 244 (50.5) | 0.02 | 0.1 |
| Private | 364 (24.9) | 141 (25.7) | 0.02 | 0 | 328 (25.4) | 121 (25.1) | -0.02 | -0.02 | 322 (25.1) | 112 (23.2) | -0.05 | -0.03 |
| Other | 63 (4.3) | 20 (3.6) | -0.02 | 0.01 | 53 (4.1) | 17 (3.5) | -0.02 | 0 | 53 (4.1) | 17 (3.5) | 0 | -0.01 |
| Unknown | 316 (21.6) | 127 (23.1) | 0.03 | 0.04 | 284 (22.0) | 110 (22.8) | 0.03 | 0.01 | 274 (21.4) | 110 (22.8) | 0.03 | 0.04 |
| Lab Parameters |  |  |  |  |  |  |  |  |  |  |  |  |
| BMI z-score | 2.3±0.6 | 2.4±0.5 | 0.17 | 0.05 | 2.3±0.6 | 2.4±0.5 | 0.15 | 0.04 | 2.3±0.6 | 2.4±0.5 | 0.15 | 0.02 |
| BMI Percentile | 97.8±4.8 | 98.3±4.2 | 0.12 | 0 | 97.7±4.9 | 98.2±4.4 | 0.1 | -0.01 | 97.8±5.0 | 98.3±4.4 | 0.11 | -0.03 |
| HbA1c <6.5 | 1255 (85.9) | 388 (70.7) | -0.38 | -0.08 | 1100 (86.0) | 340 (70.4) | -0.39 | -0.09 | 1084 (84.6) | 332 (68.7) | -0.38 | -0.15 |
| HbA1c 6.5 – 8.9 | 112 (7.7) | 70 (12.8) | 0.17 | 0.06 | 97 (7.5) | 59 (12.2) | 0.17 | 0.07 | 105 (8.2) | 61 (12.6) | 0.15 | 0.08 |
| HbA1c > 9.0 | 93 (6.4) | 90 (16.4) | 0.32 | 0.05 | 84 (6.5) | 83 (17.2) | 0.33 | 0.05 | 91 (7.1) | 89 (18.4) | 0.34 | 0.12 |
| Comorbidities |  |  |  |  |  |  |  |  |  |  |  |  |
| Type 2 Diabetes | 1345 (92.1) | 476 (86.7) | -0.17 | -0.05 | 1186 (91.9) | 414 (85.7) | -0.19 | -0.07 | 1172 (91.5) | 411 (85.1) | -0.2 | -0.14 |
| Other Diabetes | 114 (7.8) | 76 (13.8) | 0.2 | 0.05 | 103 (8.0) | 72 (14.9) | 0.22 | 0.07 | 107 (8.4) | 75 (15.5) | 0.22 | 0.14 |
| Obesity | 1221 (83.6) | 484 (88.2) | 0.13 | -0.01 | 1078 (83.5) | 424 (87.8) | 0.12 | 0 | 1065 (83.1) | 425 (88.0) | 0.14 | -0.09 |
| Overweight | 106 (7.3) | 26 (4.7) | -0.11 | 0 | 91 (7.0) | 24 (5.0) | -0.09 | -0.03 | 93 (7.3) | 22 (4.6) | -0.11 | 0.08 |
| Suicidal Ideation or Behaviors | 0 | 0 | NA | NA | ≤11 | ≤11 |  |  | 27 (2.1) | 14 (2.9) | 0.05 | -0.01 |
| Anxiety | 207 (14.2) | 80 (14.6) | 0.01 | -0.02 | 114 (8.8) | 44 (9.1) | 0.03 | 0.09 | 0 | 0 | NA | NA |
| Depression | 131 (9.0) | 54 (9.8) | 0.03 | 0 | 0 | 0 | NA | NA | 79 (6.2) | 34 (7.0) | 0.04 | -0.03 |
| Obstructive Sleep Apnea | 139 (9.5) | 77 (14.0) | 0.14 | 0.06 | 113 (8.8) | 61 (12.6) | 0.15 | 0.01 | 115 (9.0) | 60 (12.4) | 0.11 | 0.02 |
| Non-Alcoholic Fatty Liver Disease | 148 (10.1) | 91 (16.6) | 0.19 | 0.01 | 135 (10.5) | 78 (16.1) | 0.16 | -0.02 | 127 (9.9) | 75 (15.5) | 0.17 | -0.07 |
| Non-Alcoholic Steatohepatitis | 17 (1.2) | ≤11 | 0.05 | 0.05 | 16 (1.2) | ≤11 | 0.03 | 0.05 | 16 (1.2) | ≤11 | 0.06 | -0.02 |
| Anemia | 50 (3.4) | 30 (5.5) | 0.1 | -0.01 | 43 (3.3) | 22 (4.6) | 0.08 | -0.02 | 35 (2.7) | 18 (3.7) | 0.06 | -0.03 |
| Asthma | 237 (16.2) | 85 (15.5) | -0.02 | -0.03 | 205 (15.9) | 68 (14.1) | -0.04 | -0.01 | 200 (15.6) | 71 (14.7) | -0.03 | -0.03 |
| Conduct Disorders | 58 (4.0) | 14 (2.6) | -0.08 | 0.02 | 30 (2.3) | ≤11 | -0.04 | 0.04 | 43 (3.4) | 12 (2.5) | -0.05 | 0.01 |
| Nausea and Vomiting | 140 (9.6) | 54 (9.8) | 0.01 | 0.04 | 110 (8.5) | 45 (9.3) | 0.03 | 0.07 | 115 (9.0) | 39 (8.1) | -0.03 | 0.09 |
| Hypertension | 115 (7.9) | 81 (14.8) | 0.22 | -0.01 | 98 (7.6) | 66 (13.7) | 0.22 | 0.03 | 102 (8.0) | 68 (14.1) | 0.2 | 0.03 |
| Dyslipidemia | 441 (30.2) | 221 (40.3) | 0.21 | -0.01 | 403 (31.2) | 196 (40.6) | 0.19 | -0.01 | 383 (29.9) | 202 (41.8) | 0.25 | 0.01 |
| Epilepsy | 61 (4.2) | 15 (2.7) | -0.08 | 0.02 | 47 (3.6) | 10 (2.1) | -0.09 | 0.02 | 42 (3.3) | 12 (2.5) | -0.05 | -0.02 |
| Cardiovascular Conditions | 73 (5.0) | 42 (7.7) | 0.11 | 0.03 | 61 (4.7) | 34 (7.0) | 0.11 | 0.03 | 55 (4.3) | 32 (6.6) | 0.1 | 0 |
| Chromosomal Anomalies | 58 (4.0) | 20 (3.6) | -0.02 | -0.03 | 50 (3.9) | 17 (3.5) | -0.03 | 0 | 48 (3.7) | 16 (3.3) | -0.02 | -0.03 |
| Congenital Malformations | 206 (14.1) | 72 (13.1) | -0.03 | -0.01 | 178 (13.8) | 62 (12.8) | -0.03 | 0.03 | 171 (13.3) | 59 (12.2) | -0.03 | 0.02 |
| Developmental Disorders | 142 (9.7) | 40 (7.3) | -0.09 | -0.07 | 116 (9.0) | 33 (6.8) | -0.09 | -0.04 | 90 (7.0) | 33 (6.8) | -0.01 | -0.09 |
| Upper GI Conditions | 100 (6.8) | 52 (9.5) | 0.1 | -0.04 | 80 (6.2) | 39 (8.1) | 0.09 | -0.01 | 78 (6.1) | 32 (6.6) | 0.02 | -0.05 |
| Lower GI Conditions | 58 (4.0) | 33 (6.0) | 0.09 | -0.02 | 44 (3.4) | 22 (4.6) | 0.07 | 0.05 | 48 (3.7) | 21 (4.3) | 0.03 | -0.01 |
| Joint Disorders | 55 (3.8) | 21 (3.8) | 0 | -0.01 | 45 (3.5) | 18 (3.7) | 0.02 | 0.04 | 42 (3.3) | 16 (3.3) | 0 | -0.04 |
| Menstrual Disorders | 295 (20.2) | 111 (20.2) | 0 | -0.01 | 257 (19.9) | 94 (19.5) | 0 | -0.02 | 250 (19.5) | 92 (19.0) | -0.01 | -0.06 |
| Pain Conditions | 81 (5.5) | 42 (7.7) | 0.08 | 0.01 | 60 (4.6) | 30 (6.2) | 0.07 | 0.01 | 61 (4.8) | 28 (5.8) | 0.05 | -0.02 |
| Other Psychological Conditions | 55 (3.8) | 14 (2.6) | -0.07 | -0.04 | 18 (1.4) | ≤11 | -0.05 | -0.03 | 31 (2.4) | ≤11 | -0.07 | -0.05 |
| Sleep Disorders | 220 (15.1) | 99 (18.0) | 0.08 | 0 | 166 (12.9) | 73 (15.1) | 0.08 | 0 | 171 (13.3) | 72 (14.9) | 0.04 | -0.01 |
| Hearing Impairment | 35 (2.4) | 21 (3.8) | 0.08 | 0.02 | 29 (2.2) | 18 (3.7) | 0.08 | 0.03 | 27 (2.1) | 15 (3.1) | 0.06 | 0.06 |
| Polycystic Ovarian Syndrome | 245 (16.8) | 80 (14.6) | -0.06 | -0.03 | 218 (16.9) | 64 (13.3) | -0.08 | -0.04 | 213 (16.6) | 63 (13.0) | -0.1 | -0.09 |
| Thyroid Disease | 131 (9.0) | 68 (12.4) | 0.11 | -0.01 | 124 (9.6) | 57 (11.8) | 0.09 | -0.02 | 116 (9.1) | 55 (11.4) | 0.08 | -0.05 |
| Migraine | 54 (3.7) | 25 (4.6) | 0.04 | -0.03 | 39 (3.0) | 14 (2.9) | 0 | 0.04 | 33 (2.6) | 12 (2.5) | -0.01 | 0.02 |
| Hyperglycemic Emergency | 29 (2.0) | 19 (3.5) | 0.09 | 0.07 | 25 (1.9) | 19 (3.9) | 0.12 | 0.06 | 29 (2.3) | 14 (2.9) | 0.1 | 0.14 |
| Hypoglycemia Crisis | 50 (3.4) | 29 (5.3) | 0.09 | -0.01 | 47 (3.6) | 26 (5.4) | 0.08 | 0 | 27 (2.1) | 18 (3.7) | 0.1 | 0.05 |
| Comedications |  |  |  |  |  |  |  |  |  |  |  |  |
| Insulin | 178 (12.2) | 156 (28.4) | 0.41 | 0.06 | 161 (12.5) | 143 (29.6) | 0.43 | 0.06 | 168 (13.1) | 150 (31.1) | 0.44 | 0.13 |
| Sodium Glucose Co-transporter 2 inhibitors | ≤11 | ≤11 | 0.07 | 0 | ≤11 | ≤11 | 0.06 | 0.01 | ≤11 | ≤11 | 0.06 | 0.02 |
| Bupropion | ≤11 | ≤11 | 0.09 | 0 | ≤11 | ≤11 | 0.08 | 0.01 | ≤11 | ≤11 | 0.02 | -0.06 |
| Orlistat | ≤11 | ≤11 | 0.08 | 0.02 | ≤11 | ≤11 | 0.09 | 0.01 | ≤11 | ≤11 | 0.03 | 0.01 |
| ACE Inhibitors | 38 (2.6) | 33 (6.0) | 0.17 | 0.01 | 31 (2.4) | 29 (6.0) | 0.18 | 0 | 34 (2.7) | 30 (6.2) | 0.17 | 0.01 |
| Beta Blockers | 16 (1.1) | 12 (2.2) | 0.09 | -0.03 | ≤11 | ≤11 | 0.09 | -0.02 | 17 (1.3) | ≤11 | 0.03 | -0.01 |
| Calcium Channel Blockers | 16 (1.1) | 13 (2.4) | 0.1 | -0.01 | ≤11 | 12 (2.5) | 0.12 | -0.01 | 14 (1.1) | ≤11 | 0.08 | 0.12 |
| Diuretics | 36 (2.5) | 19 (3.5) | 0.06 | -0.02 | 32 (2.5) | 14 (2.9) | 0.05 | -0.02 | 31 (2.4) | 15 (3.1) | 0.04 | -0.02 |
| ARBs | 12 (0.8) | 13 (2.4) | 0.12 | -0.05 | ≤11 | ≤11 | 0.13 | -0.03 | 12 (0.9) | ≤11 | 0.06 | -0.04 |
| Statin Use | 32 (2.2) | 32 (5.8) | 0.19 | -0.08 | 28 (2.2) | 29 (6.0) | 0.19 | -0.08 | 29 (2.3) | 29 (6.0) | 0.19 | -0.05 |
| Other Lipid-lowering Agents | ≤11 | 13 (2.4) | 0.14 | -0.01 | ≤11 | ≤11 | 0.14 | 0 | ≤11 | ≤11 | 0.14 | -0.03 |
| Antidepressants | 59 (4.0) | 26 (4.7) | 0.03 | -0.08 | 0 | 0 | NA | NA | 35 (2.7) | 12 (2.5) | -0.02 | -0.05 |
| Antipsychotics | 120 (8.2) | 17 (3.1) | -0.22 | -0.04 | 51 (4.0) | 9 (1.9) | -0.19 | -0.01 | 79 (6.2) | ≤11 | -0.19 | -0.05 |
| Benzodiazepines | 82 (5.6) | 35 (6.4) | 0.03 | 0.01 | 58 (4.5) | 25 (5.2) | 0.02 | 0.01 | 70 (5.5) | 24 (5.0) | -0.02 | -0.05 |
| Proton Pump Inhibitors | 68 (4.7) | 42 (7.7) | 0.12 | -0.01 | 52 (4.0) | 25 (5.2) | 0.1 | -0.02 | 55 (4.3) | 24 (5.0) | 0.03 | -0.03 |
| Oral Steroids | 318 (21.8) | 108 (19.7) | -0.05 | 0 | 265 (20.5) | 80 (16.6) | -0.08 | 0 | 268 (20.9) | 84 (17.4) | -0.09 | 0.01 |
| Opioids | 84 (5.7) | 45 (8.2) | 0.1 | -0.02 | 65 (5.0) | 34 (7.0) | 0.11 | -0.01 | 68 (5.3) | 34 (7.0) | 0.07 | -0.04 |
| NSAIDs | 182 (12.5) | 94 (17.1) | 0.13 | -0.02 | 143 (11.1) | 70 (14.5) | 0.11 | 0.03 | 151 (11.8) | 69 (14.3) | 0.07 | -0.03 |
| Direct Oral Anticoagulants | ≤11 | ≤11 | 0.04 | 0.03 | ≤11 | ≤11 | 0.06 | 0.02 | ≤11 | ≤11 | 0.05 | 0 |
| Provider Visits |  |  |  |  |  |  |  |  |  |  |  |  |
| Pediatrician Visit | 1165 (79.7) | 421 (76.7) | -0.07 | 0.03 | 1048 (81.2) | 373 (77.2) | -0.07 | 0.02 | 1025 (80.0) | 372 (77.0) | -0.07 | -0.01 |
| Primary Care Visit | 68 (4.7) | 18 (3.3) | -0.07 | 0.06 | 58 (4.5) | 14 (2.9) | -0.09 | 0.02 | 59 (4.6) | 14 (2.9) | -0.09 | 0.13 |
| Psychiatric Visit | 152 (10.4) | 33 (6.0) | -0.16 | -0.05 | 72 (5.6) | 19 (3.9) | -0.09 | 0.07 | 83 (6.5) | 20 (4.1) | -0.1 | -0.08 |
| Counseling Visit | 74 (5.1) | 32 (5.8) | 0.03 | 0.06 | 46 (3.6) | 19 (3.9) | -0.1 | -0.01 | 38 (3.0) | 20 (4.1) | 0.06 | 0.01 |
| Nutritionist Visit | 401 (27.4) | 209 (38.1) | 0.23 | 0.03 | 353 (27.3) | 178 (36.9) | 0.02 | 0.04 | 328 (25.6) | 179 (37.1) | 0.25 | -0.03 |
| Childhood Comorbidity Index |  |  |  |  |  |  |  |  |  |  |  |  |
| Childhood Comorbidity Index < 4 | 781 (53.5) | 164 (29.9) | -0.49 | -0.14 | 765 (59.3) | 164 (34.0) | -0.52 | -0.16 | 736 (57.5) | 157 (32.5) | -0.52 | -0.18 |
| Childhood Comorbidity Index ≥ 4 | 680 (46.5) | 385 (70.1) |  |  | 526 (40.7) | 319 (66.0) |  |  | 545 (42.5) | 326 (67.5) |  |  |

To evaluate covariate balance before and after weighting, standardized mean differences (SMDs) were calculated, and values below 0.1 were considered indicative of negligible imbalance between treatment groups.^29^ We followed patients from cohort entry until the earliest of the following: 1) incidence of the study outcome, 2) death, or 3) end of follow-up, i.e., the last documented encounter in the OneFlorida+ network or January 31, 2024, whichever occurred first.

The outcome comparisons between GLP-1RA and prevalent metformin users were conducted using sIPTW-weighted Cox proportional hazards models to estimate hazard ratios (HRs) and 95% confidence intervals (CIs). Variables with an aSMD >0.1 were re-adjusted in the Cox model. A *p*-value <0.05 was considered statistically significant. Additionally, we estimated the incidence per 1000-person years for each outcome and calculated the rate difference (RD) between groups. Additionally,

We performed subgroup analyses by YT2D status, age (<12 vs. ≥12 years), race/ethnicity, and sex. We also carried out as-treated sensitivity analyses where we applied a 90-day and 60-day grace period to bridge consecutive prescriptions that did not overlap.

All statistical analyses were performed using SAS, version 9.4 (SAS Institute Inc., Cary, NC, USA).

## RESULTS

The flowchart of patient selection based on the inclusion and exclusion criteria is presented in **Figure 1**. We included 2,010 patients in the cohort to examine incident suicidal ideation or behaviors (549 GLP-1 RA vs 1,461 prevalent metformin users), 1,774 patients in the cohort to assess depression outcome (483 GLP-1 RA vs 1,291 prevalent metformin users), and 1,764 patients in the cohort to assess incident anxiety (483 GLP-1 RA vs 1,281 prevalent metformin users). For the cohort evaluating suicide ideation and behaviors, GLP-1RA initiators were generally older (mean ± SD age: 14.7±2.1 vs 13.7±2.5 years), non-Hispanic Black (28.4% vs 26.4%), with private insurance (25.7% vs 24.9%), a higher mean baseline BMI-z score (2.4±0.5 vs 2.3±0.6), BMI percentile (98.3±4.2 vs 97.8±4.8) and more likely to have an HbA1c ≥6.5% (29.2% vs 14.1%) compared with prevalent metformin users. The cohorts evaluating depression and anxiety showed similar trends in sociodemographic characteristics. Additionally, these two cohorts had a higher proportion of Hispanic GLP-1 RA users compared to prevalent metformin users (40.9% vs 39.8% and 42% vs 40.9%, respectively). The baseline characteristics of patients in each of these cohorts are presented in **Table 1**.

**Figure 1:**
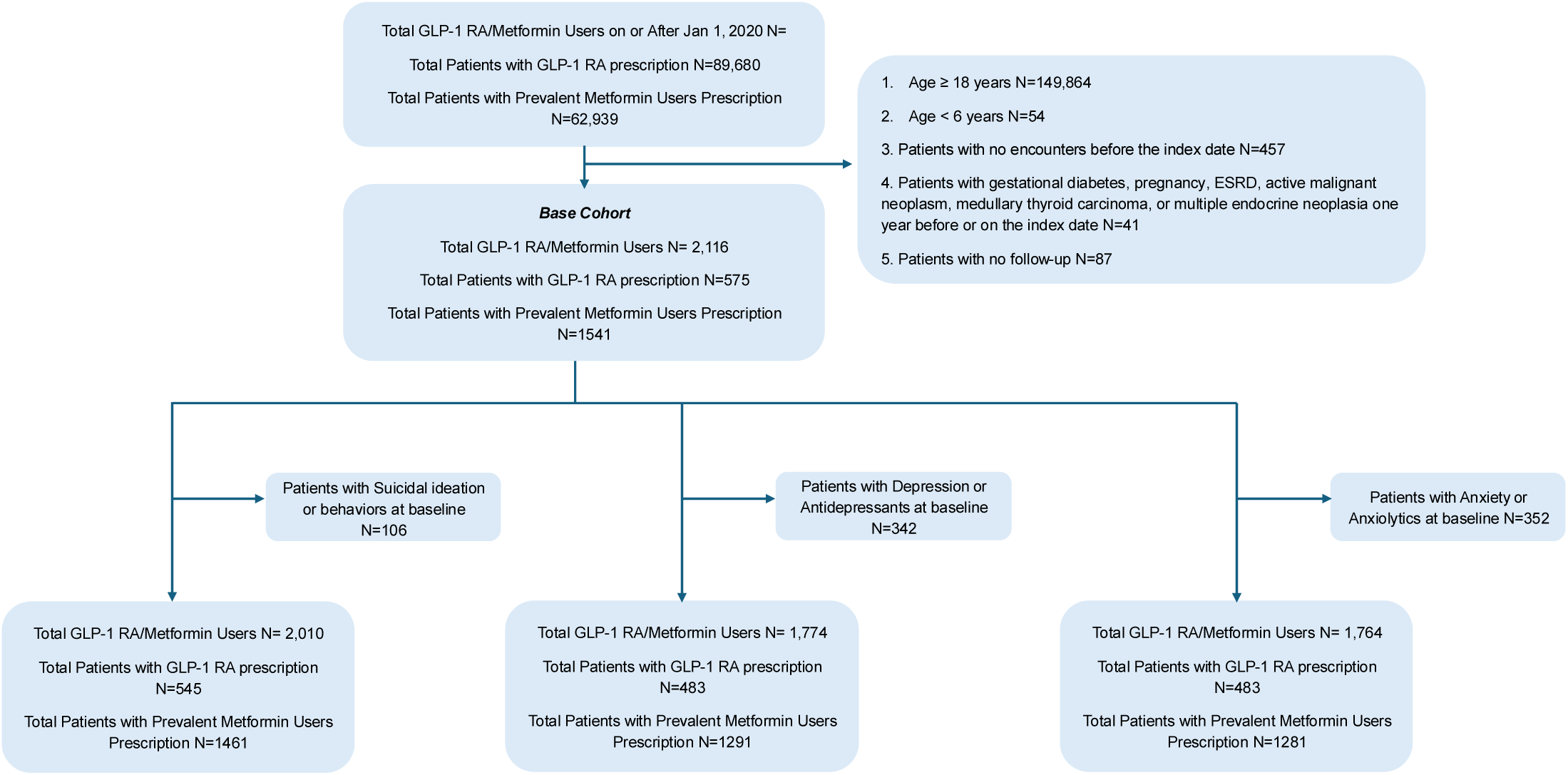
Cohort Selection

For the outcome of suicidal ideation and behaviors, ≤11 cases were identified among GLP-1RA users (median, interquartile range (IQR) follow-up: 1.51 years, 0.76-2.59 years) compared with 28 cases (2.1%) among prevalent metformin users (median, IQR follow-up: 1.48 years, 0.70-2.53 years), during follow-up. The incidence rate for suicidal ideation and behaviors was 1.50 and 11.94 per 1000 person-years for the GLP-1RA and the prevalent metformin groups, respectively, yielding an RD of -10.45 (95% CI, -14.54 to -6.36) per 1000 person-years. GLP-1RA use was statistically significantly associated with a decreased risk of suicidal ideation or behaviors, with an adjusted HR of 0.11 (95% CI, 0.02 to 0.86). (**Figure 2**) The results were consistent across subgroup analyses. (**Figure 3**)

**Figure 2:**
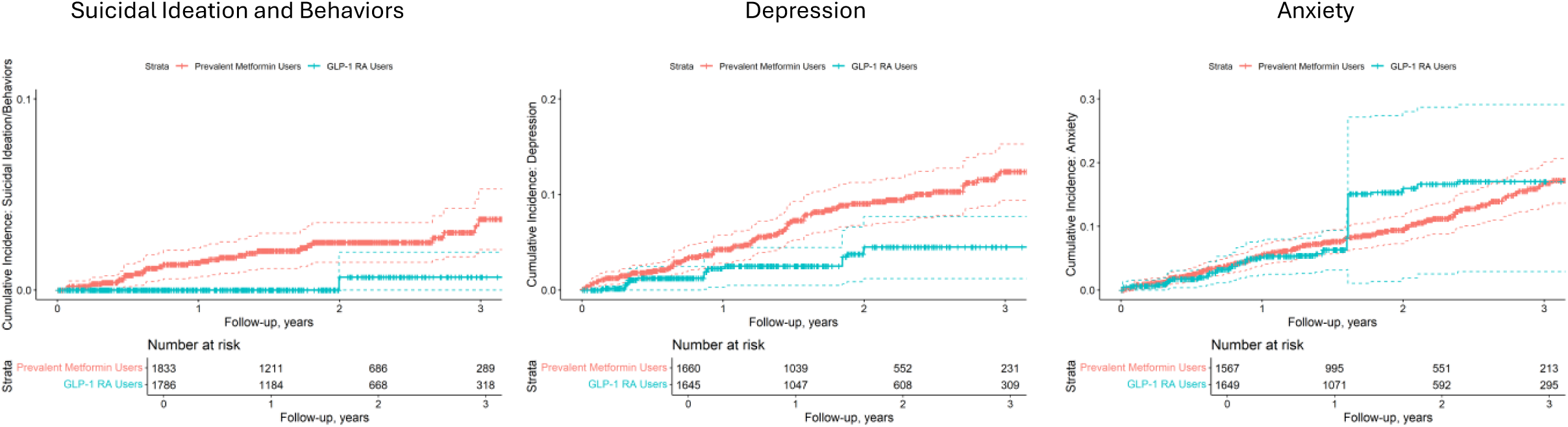
Inverse Probability of Treatment Weighting (IPTW)–Adjusted Cumulative Incidence Within the 3 Study Cohorts

**Figure 3:**
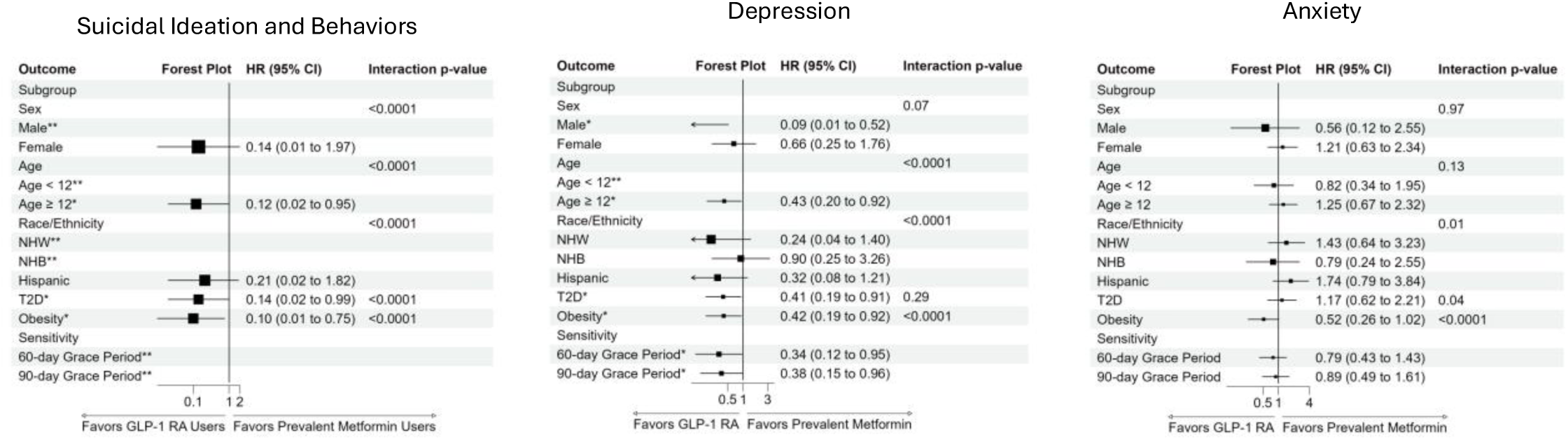
Subgroup Analysis: Inverse Probability of Treatment Weighting (IPTW) Adjusted Hazard Ratio for Each Outcome by Subgroup *Indicates significance for Hazard Ratio **HR (95% CI) could not be estimated due to no events in one of the groups upon re-weighting for subgroup analysis or during the mentioned period for sensitivity analysis. *Abbreviations*: T2D: Type 2 Diabetes, NHW: Non-Hispanic White, NHB: Non-Hispanic Black

For the outcome of depression, ≤11 cases were identified among GLP-1RA users (median, IQR follow-up: 1.43 years, 0.72-2.61 years) compared with 79 cases (6.9%) among prevalent metformin users (median, IQR follow-up: 1.36 years, 0.65-2.40 years), during follow-up. The incidence rate for depression was 16.95 and 42.59 per 1000 person-years for the GLP-1RA and the prevalent metformin groups, respectively, yielding an RD of -25.64 (95% CI, -34.90 to -16.39) per 1000 person-years. GLP-1RA use was statistically significantly associated with a decreased risk of depression, with an adjusted HR of 0.37 (95% CI, 0.17 to 0.78). (**Figure 2**) The results were consistent across subgroups and sensitivity analyses depicting a lower risk of depression in GLP-1 RA users (**Figure 3**).

For the outcome of anxiety, 30 (6.8%) cases were identified among GLP-1RA users (median, IQR follow-up: 1.56 years, 0.74-2.55 years) compared with 110 incident cases (9.7%) among prevalent metformin users (median, IQR follow-up: 1.40 years, 0.67-2.39 years), during follow-up. The incidence rate for anxiety was 61.49 and 55.54 per 1000 person-years for the GLP-1RA and the prevalent metformin groups, respectively, yielding an RD of 5.95 (95% CI, - 7.10 to 19.01) per 1000 person-years. GLP-1RA use was not significantly associated with the risk of anxiety, with an adjusted HR of 1.13 (95% CI, 0.69 to 1.84). (**Figure 2**) The results were consistent across subgroups and sensitivity analyses, with no significant difference between the two groups. (**Figure 3**)

## DISCUSSION

In this target trial emulation study that includes a real-world cohort of children and adolescents with obesity or YT2D, we observed that, in the short-term (might want to explain mean f/u time here),the use of GLP-1 RA was associated with a lower risk of suicidal attempts and depression compared to prevalent metformin users. The risk of anxiety was not significantly different between the two groups. These findings provide real-world evidence supporting the mental health safety profile of GLP-1 RAs in pediatric populations. However, surveillance is needed over a longer follow-up

We observed that the risk of suicidal ideation and behaviors was significantly lower in children and adolescents using GLP-1 RA vs prevalent metformin users. Our result was in line with a recent study by Kerem et al, using TriNetX data, found that the risk of suicidal ideation or behaviors was lowered in children and adolescents with obesity using GLP-1 RAs vs lifestyle intervention.^30^ While we saw consistent lowering trends in suicidal ideation or behaviors by subgroup, some subgroup analyses revealed contrasting findings, such as we found a lower risk of suicidal ideation or behaviors in the male and YT2D subgroups, whereas Kerem et. al found no significant differences for these subgroups, and vice-versa for the female subgroups.^30^ Interestingly, while the findings from the main analysis remained consistent with our study, the differences in subgroup analysis could reveal the differences in study design, i.e., we included an expanded population of children and adolescents with obesity or YT2D (vs. obesity alone) and used a clinically relevant comparator like prevalent metformin users (vs. lifestyle intervention which is a first-line treatment before pharmacotherapy). However, these results in children and adolescents seem to contrast with the adult population.^13,14^ This could be attributed to the shorter duration of follow-up due to shorter time on market for pediatric populations compared to adults, additional caution on the physician’s end, or possibly due to the variability in pharmacokinetics and pharmacodynamics of medications in pediatrics vs adults.^31^ Continued surveillance with longer follow-up is needed in this population before the protective effects can be confirmed.

Additionally, GLP-1 RA use was also associated with a lower risk of depression compared to prevalent metformin users, with consistent trends observed in subgroups. Real-world evidence on GLP-1 RA use and depression or anxiety risk in children and adolescents is lacking. However, studies in adults show lower depression risk with GLP-1 RAs compared to dipeptidyl peptidase-4 inhibitor users, but no significant difference compared to sodium-glucose cotransporter-2 inhibitor users.^32^ A recent meta-analysis of over 2,000 adults found GLP-1 RAs significantly reduced depression scores, aligning with our findings.^33^ In contrast, we found a non-significant but increasing trend in anxiety risk between the treatment groups, and subgroup trends remained non-significant and inconsistent. A study by Lu et al. using the FDA Adverse Event Reporting System database found weak signals for anxiety with GLP-1 RA use in adults, consistent with our findings.^34^

Overall, the potential antidepressant and anxiogenic effects of GLP-1 RAs align with biological mechanisms observed in preclinical studies.^35,36^ Anxiogenic effects are linked to dorsal raphe activation, which increases anxiety-like behavior via heightened serotonergic transmission in the amygdala. In contrast, antidepressant effects may result from reduced neuroinflammation, enhanced neurogenesis, and modulation of brain activity.^35^ These studies demonstrated that GLP-1 RAs induce anxiety after initiating treatment, however, it seemed to be replaced by positive effects on mood, manifested by reduced depression-like behavior.^35^ These results may have direct clinical relevance as they warrant vigilance on the part of treating pediatricians during the initial phase of the treatment, as elevated anxiety may be observed in the initial treatment phase, similar to selective serotonin reuptake inhibitors and indicate that GLP-1 RA may be useful for patients with YT2D and obesity manifesting with comorbid depression.

This study has several strengths. First, to the best of our knowledge, this is the first real-world study exploring GLP-1 RA use and the risk of psychological outcomes and behaviors in children and adolescents with YT2D and/or obesity. Additionally, despite concerns by regulatory authorities around the world, including the U.S. FDA and EMA, regarding GLP-1 RA use and suicidal ideation and behaviors, the real-world evidence is largely limited to adults, with little to no focus on this growing young population. Second, using a clinically relevant comparator like prevalent metformin users is helpful from a clinical perspective to guide certain decisions and look out for any potential signals in practice. Third, using EHR data like OneFLorida+ makes the data rich in insurance representativeness from private to Medicaid and no insurance groups, and helps account for clinical confounders like BMI, BMI-z score, BMI percentiles, and HbA1c. Lastly, and importantly, our results may have clinical implications, such as an increase in active surveillance from treating pediatricians and psychologists due to the effect of GLP-1 RAs on biological mechanisms to mimic anxiogenic effects.

However, our study is subject to limitations, and the findings must be interpreted with caution. First, there is a possibility of prescriptions outside the OneFlorida+ network, which cannot be captured in the current EHR data. Second, some limitations inherent to observational studies, including potential unmeasured confounding due to the lack of information about family history and the patient’s duration of diabetes, and obesity. Lastly, as GLP-1 RAs are relatively newer generation pharmacologic agents approved among pediatric populations, we were limited by sample size and relatively short follow-up, leading to relatively wide confidence intervals and limiting certain subgroup analyses by outcome.

In conclusion, in the short term, GLP-1 RAs may be associated with a lower risk of suicidal ideation and behaviors, as well as depression, with no evidence of an association with heightened anxiety risk in children and adolescents with obesity or YT2D. These findings suggest a relatively favorable mental health safety profile for GLP-1 RAs in children and adolescents. Future studies with larger sample sizes and longer follow-up periods are warranted to confirm and extend these observations.

## Data Availability

Data set Available through OneFlorida+ Clinical Research Network (email)

## REFERENCES

1. Hampl SE, Hassink SG, Skinner AC, et al. Clinical Practice Guideline for the Evaluation and Treatment of Children and Adolescents With Obesity. Pediatrics. Feb 01 2023;151(2).

2. Kansra AR, Lakkunarajah S, Jay MS. Childhood and Adolescent Obesity: A Review. Front Pediatr. 2020;8:581461.

3. Centers for Disease Prevention and Control. Diabetes. 2024.

4. Merwass NA, Alkhader YK, Alharthi SA, et al. The Role of Screening, Risk Factors, and Early Intervention in Preventing Diabetes in the Obese Population: A Systematic Review. Cureus. Jul 2024;16(7):e63952.

5. Shaban Mohamed MA, AbouKhatwa MM, Saifullah AA, et al. Risk Factors, Clinical Consequences, Prevention, and Treatment of Childhood Obesity. Children (Basel). Dec 16 2022;9(12).

6. Borzutzky C, King E, Fox CK, et al. Trends in prescribing anti-obesity pharmacotherapy for paediatric weight management: Data from the POWER Work Group. Pediatr Obes. Jan 2021;16(1):e12701.

7. Raman V, Foster CM. Metformin Treatment of Pediatric Obesity. Pediatrics. Mar 2021;147(3).

8. Lee JM, Sharifi M, Oshman L, Griauzde DH, Chua KP. Dispensing of Glucagon-Like Peptide-1 Receptor Agonists to Adolescents and Young Adults, 2020-2023. JAMA. Jun 18 2024;331(23):2041–2043.

9. Committee ADAPP. 14. Children and Adolescents: Standards of Care in Diabetes-2024. Diabetes Care. Jan 01 2024;47(Suppl 1):S258–S281.

10. ClinicalTrials.gov. A Study of Tirzepatide (LY3298176) Once Weekly in Adolescent Participants Who Have Obesity or Overweight With Weight-Related Comorbidities.

11. European Medicines Agency. EMA statement on ongoing review of GLP-1 receptor agonists. 2023.

12. Food and Drug Administration. Update on FDA’s ongoing evaluation of reports of suicidal thoughts or actions in patients taking a certain type of medicines approved for type 2 diabetes and obesity. 2024.

13. Ueda P, Söderling J, Wintzell V, et al. GLP-1 Receptor Agonist Use and Risk of Suicide Death. JAMA Intern Med. Sep 03 2024.

14. Tang H, Lu Y, Donahoo WT, et al. Glucagon-Like Peptide-1 Receptor Agonists and Risk for Suicidal Ideation and Behaviors in U.S. Older Adults With Type 2 Diabetes : A Target Trial Emulation Study. Ann Intern Med. Aug 2024;177(8):1004–1015.

15. McIntyre RS, Mansur RB, Rosenblat JD, Kwan ATH. The association between glucagon-like peptide-1 receptor agonists (GLP-1 RAs) and suicidality: reports to the Food and Drug Administration Adverse Event Reporting System (FAERS). Expert Opin Drug Saf. Jan 2024;23(1):47–55.

16. American Academy of Pediatrics. Suicide: Pediatric Mental Health Minute Series.

17. Bitsko RH, Claussen AH, Lichstein J, et al. Mental Health Surveillance Among Children - United States, 2013-2019. MMWR Suppl. Feb 25 2022;71(2):1–42.

18. U.S. Centers for Disease Control and Prevention. Children’s Mental Health. 2025.

19. Hogan WR, Shenkman EA, Robinson T, et al. The OneFlorida Data Trust: a centralized, translational research data infrastructure of statewide scope. J Am Med Inform Assoc. Mar 15 2022;29(4):686–693.

20. Center for Drug Evaluation and Research. Guidance for Industry: Suicidal Ideation and Behavior: Prospective Assessment of Occurrence in Clinical Trials. 2012.

21. Agency for Healthcare Research and Quality. Suicidal Ideation, Suicide Attempt, or Self-Inflicted Harm: Pediatric Emergency Department Visits, 2010–2014 and 2016. Healthcare Cost and Utilization Project2019.

22. Swain RS, Taylor LG, Braver ER, Liu W, Pinheiro SP, Mosholder AD. A systematic review of validated suicide outcome classification in observational studies. Int J Epidemiol. Oct 01 2019;48(5):1636–1649.

23. Hedegaard H, Schoenbaum M, Claassen C, Crosby A, Holland K, Proescholdbell S. Issues in Developing a Surveillance Case Definition for Nonfatal Suicide Attempt and Intentional Self-harm Using International Classification of Diseases, Tenth Revision, Clinical Modification (ICD-10-CM) Coded Data. Natl Health Stat Report. Feb 2018(108):1–19.

24. Schein J, Childress A, Gagnon-Sanschagrin P, et al. Treatment Patterns Among Patients with Attention-Deficit/Hyperactivity Disorder and Comorbid Anxiety and/or Depression in the United States: A Retrospective Claims Analysis. Adv Ther. May 2023;40(5):2265–2281.

25. American Psychiatric Association. Diagnostic and statistical manual of mental disorders, 5th Edition. 2013.

26. U.S. Centers for Disease Control and Prevention. SAS Program for CDC Growth Charts. 2024.

27. Sun JW, Bourgeois FT, Haneuse S, et al. Development and Validation of a Pediatric Comorbidity Index. Am J Epidemiol. May 04 2021;190(5):918–927.

28. Xu S, Ross C, Raebel MA, Shetterly S, Blanchette C, Smith D. Use of stabilized inverse propensity scores as weights to directly estimate relative risk and its confidence intervals. Value Health. 2010;13(2):273–277.

29. Franklin JM, Rassen JA, Ackermann D, Bartels DB, Schneeweiss S. Metrics for covariate balance in cohort studies of causal effects. Stat Med. May 10 2014;33(10):1685–1699.

30. Kerem L, Stokar J. Risk of Suicidal Ideation or Attempts in Adolescents With Obesity Treated With GLP1 Receptor Agonists. JAMA Pediatr. Dec 01 2024;178(12):1307–1315.

31. Stephenson T. How children’s responses to drugs differ from adults. Br J Clin Pharmacol. Jun 2005;59(6):670–673.

32. Tang H, Lu Y, Donahoo WT, et al. Glucagon-Like Peptide-1 Receptor Agonists and Risk for Depression in Older Adults With Type 2 Diabetes : A Target Trial Emulation Study. Ann Intern Med. Mar 2025;178(3):315–326.

33. Chen X, Zhao P, Wang W, Guo L, Pan Q. The Antidepressant Effects of GLP-1 Receptor Agonists: A Systematic Review and Meta-Analysis. Am J Geriatr Psychiatry. Jan 2024;32(1):117–127.

34. Lu W, Wang S, Tang H, Yuan T, Zuo W, Liu Y. Neuropsychiatric adverse events associated with Glucagon-like peptide-1 receptor agonists: a pharmacovigilance analysis of the FDA Adverse Event Reporting System database. Eur Psychiatry. Feb 04 2025;68(1):e20.

35. Anderberg RH, Richard JE, Hansson C, Nissbrandt H, Bergquist F, Skibicka KP. GLP-1 is both anxiogenic and antidepressant; divergent effects of acute and chronic GLP-1 on emotionality. Psychoneuroendocrinology. Mar 2016;65:54–66.

36. Kim YK, Kim OY, Song J. Alleviation of Depression by Glucagon-Like Peptide 1 Through the Regulation of Neuroinflammation, Neurotransmitters, Neurogenesis, and Synaptic Function. Front Pharmacol. 2020;11:1270.

