## Supplement 1 for "Risk of Suicidal Ideation and Behaviors, Depression, and Anxiety with GLP-1 Receptor Agonist Use in Children and Adolescents: A Target Trial Emulation Study"

### Supplemental Online Content 1

**eTable 1: Target Trial Emulation Framework**

**eTable 2: International Classification of Diseases (ICD) Codes for the Outcomes**

**eTable 3: Medication Codes for GLP-1 Receptor Agonists**

**eTable 1: Target Trial Emulation Framework**

| Approaches | Target Trial | Target Trial Emulation |
|---|---|---|
| **Eligibility Criteria** | Target trial among children and adolescents with obesity or type 2 diabetes<br>• Age≥ 6 and <18 years between 1 January 2020 and 31 January 2024<br>• Type 2 diabetes and Obesity (ascertained using labs and diagnosis codes)<br>• No previous prescriptions for GLP-1RA in the past year<br>• Present at the screening<br>• No diagnosis of gestational diabetes<br>• Not pregnant<br>• No active neoplasm or thyroid medullary carcinoma<br>• No diagnosis of ESRD<br>• No history of suicidal ideation or suicidal behaviors/depression / anxiety, depending on the outcome being evaluated | Same as the target trial |
| **Treatment Strategies** | GLP-1 RA use vs. Placebo | GLP-1 RA use vs. Prevalent Metformin User |
| **Treatment assignment** | Individuals are randomly assigned to a treatment at baseline.<br>Individuals and their treating physicians will be aware of the assigned treatment strategy. | The date of medication initiation/ the follow-up start date was defined as:<br>• The first date of GLP-1 RA prescription for new users<br>• The consecutive prescription fill of metformin between 2020-2024 for prevalent metformin users<br>Stabilized inverse probability of treatment weighting (sIPTW) |
| **Outcomes** | Suicidal Ideation and Behaviors, Depression and Anxiety, prospective surveillance during efficacy trials | Suicidal Ideation and Behaviors, Depression and Anxiety, measured via diagnosis codes, recorded in the database prospectively, but collected by researchers retrospectively. |
| **Follow-up** | Follow-up starts on the treatment assignment date and is carried out until death or the end of the trial period. | Follow patients from cohort entry until the earliest of the following: 1) Study Outcome, or 2) death, or 3) end of follow-up, i.e., the last documented encounter |

| | | |
|---|---|---|
| | | in the OneFlorida+ network or January 31, 2024, whichever occurs first |
| **Causal Contrasts** | Intention-to-treat effect and per-protocol effect | Same as the target trial |
| **Statistical Analysis** | Cox proportional hazards regression model to estimate hazard ratios and Kaplan-Meier plot to show the cumulative incidence over time.<br>Intention-to-treat analysis;<br>Sensitivity analyses: per-protocol analysis;<br>(<12 and ≥12 years), sex (male and female), race, T2D or no T2D | Same as the target trial. |

**eTable 2: International Classification of Diseases (ICD) Codes for the Outcomes**

| Condition | ICD-9 | ICD-10 |
|---|---|---|
| **Obesity** | | |
| Obesity | 278.00, 278.01, V85.54 | E66.811, E66.812, E66.813, E66.01, E66.09, E66.89, E66.9, E66.0, Z68.54, Z68.55, Z68.56 or Body Mass Index percentile ≥95 % |
| **Type 2 Diabetes** | | |
| Type 2 Diabetes | 250.x0, 250.x2 | E11.xx |
| **Suicidal Ideation and Behaviors** | | |
| **Self-Harm and Suicidal Behaviors** | E950.0, E950.1, E950.2, E950.3, E950.4, E950.5, E950.6, E950.7, E950.8, E950.9, E951.0, E951.1, E951.8, E952.0, E952.1, E952.8, E952.9, E953.0, E953.1, E953.8, E953.9, E954, E955.0, E955.1, E955.2, E955.3, E955.4, E955.5, E955.6, E955.7, E955.9, E956, E957.0, E957.1, E957.2, E957.9, E958.0, E958.1, E958.2, E958.3, E958.4, E958.5, E958.6, E958.7, E958.8, E958.9 | T36.0X2A, T36.1X2A, T36.2X2A, T36.3X2A, T36.4X2A, T36.5X2A, T36.6X2A, T36.7X2A, T36.8X2A, T36.92XA, T37.0X2A, T37.1X2A, T37.2X2A, T37.3X2A, T37.4X2A, T37.5X2A, T37.8X2A, T37.92XA, T38.0X2A, T38.1X2A, T38.2X2A, T38.3X2A, T38.4X2A, T38.5X2A, T38.6X2A, T38.7X2A, T38.802A, T38.812A, T38.892A, T38.902A, T38.992A, T39.012A, T39.092A, T39.1X2A, T39.2X2A, T39.312A, T39.392A, T39.4X2A, T39.8X2A, T39.92XA, T40.0X2A, T40.1X2A, T40.2X2A, T40.3X2A, T40.4X2A, T40.5X2A, T40.602A, T40.692A, T40.7X2A, T40.8X2A, T40.902A, T40.992A, T41.0X2A, T41.1X2A, T41.202A, T41.292A, T41.3X2A, T41.42XA, T41.5X2A, T42.0X2A, T42.1X2A, T42.2X2A, T42.3X2A, T42.4X2A, T42.5X2A, T42.6X2A, T42.72XA, T42.8X2A, T43.012A, T43.022A, T43.1X2A, T43.202A, T43.212A, T43.222A, T43.292A, T43.3X2A, T43.4X2A, T43.502A, T43.592A, T43.602A, T43.612A, T43.622A, T43.632A, T43.692A, T43.8X2A, T43.92XA, T44.0X2A, |

| | | |
|---|---|---|
| | | T44.1X2A, T44.2X2A, T44.3X2A, T44.4X2A, T44.5X2A, T44.6X2A, T44.7X2A, T44.8X2A, T44.902A, T44.992A, T45.0X2A, T45.1X2A, T45.2X2A, T45.3X2A, T45.4X2A, T45.512A, T45.522A, T45.602A, T45.612A, T45.622A, T45.692A, T45.7X2A, T45.8X2A, T45.92XA, T46.0X2A, T46.1X2A, T46.2X2A, T46.3X2A, T46.4X2A, T46.5X2A, T46.6X2A, T46.7X2A, T46.8X2A, T46.902A, T46.992A, T47.0X2A, T47.1X2A, T47.2X2A, T47.3X2A, T47.4X2A, T47.5X2A, T47.6X2A, T47.7X2A, T47.8X2A, T47.92XA, T48.0X2A, T48.1X2A, T48.202A, T48.292A, T48.3X2A, T48.4X2A, T48.5X2A, T48.6X2A, T48.902A, T48.992A, T49.0X2A, T49.1X2A, T49.2X2A, T49.3X2A, T49.4X2A, T49.5X2A, T49.6X2A, T49.7X2A, T49.8X2A, T49.92XA, T50.0X2A, T50.1X2A, T50.2X2A, T50.3X2A, T50.4X2A, T50.5X2A, T50.6X2A, T50.7X2A, T50.8X2A, T50.902A, T50.992A, T50.A12A, T50.A22A, T50.A92A, T50.B12A, T50.B92A, T50.Z12A, T50.Z92A, T51.0X2A, T51.1X2A, T51.2X2A, T51.3X2A, T51.8X2A, T51.92XA, T52.0X2A, T52.1X2A, T52.2X2A, T52.3X2A, T52.4X2A, T52.8X2A, T52.92XA, T53.0X2A, T53.1X2A, T53.2X2A, T53.3X2A, T53.4X2A, T53.5X2A, T53.6X2A, T53.7X2A, T53.92XA, T54.0X2A, T54.1X2A, T54.2X2A, T54.3X2A, |

| | | |
|---|---|---|
| | | T54.92XA, T55.0X2A, T55.1X2A, T56.0X2A, T56.1X2A, T56.2X2A, T56.3X2A, T56.4X2A, T56.5X2A, T56.6X2A, T56.7X2A, T56.812A, T56.892A, T56.92XA, T57.0X2A, T57.1X2A, T57.2X2A, T57.3X2A, T57.8X2A, T57.92XA, T58.02XA, T58.12XA, T58.2X2A, T58.8X2A, T58.92XA, T59.0X2A, T59.1X2A, T59.2X2A, T59.3X2A, T59.4X2A, T59.5X2A, T59.6X2A, T59.7X2A, T59.812A, T59.892A, T59.92XA, T60.0X2A, T60.1X2A, T60.2X2A, T60.3X2A, T60.4X2A, T60.8X2A, T60.92XA, T61.02XA, T61.12XA, T61.772A, T61.782A, T61.8X2A, T61.92XA, T62.0X2A, T62.1X2A, T62.2X2A, T62.8X2A, T62.92XA, T63.002A, T63.012A, T63.022A, T63.032A, T63.042A, T63.062A, T63.072A, T63.082A, T63.092A, T63.112A, T63.122A, T63.192A, T63.2X2A, T63.302A, T63.312A, T63.322A, T63.332A, T63.392A, T63.412A, T63.422A, T63.432A, T63.442A, T63.452A, T63.462A, T63.482A, T63.512A, T63.592A, T63.612A, T63.622A, T63.632A, T63.692A, T63.712A, T63.792A, T63.812A, T63.822A, T63.832A, T63.892A, T63.92XA, T64.02XA, T64.82XA, T65.0X2A, T65.1X2A, T65.212A, T65.222A, T65.292A, T65.3X2A, T65.4X2A, T65.5X2A, T65.6X2A, T65.812A, T65.822A, T65.832A, T65.892A, T65.92XA, T71.112A, T71.122A, T71.132A, T71.152A, T71.162A, T71.192A, T71.222A, T71.232A, X71.0XXA, X71.1XXA, X71.2XXA, X71.3XXA, X71.8XXA, X71.9XXA, X72.XXXA, |

| | | |
|---|---|---|
| | | X73.0XXA, X73.1XXA, X73.2XXA, X73.8XXA, X73.9XXA, X74.01XA, X74.02XA, X74.09XA, X74.8XXA, X74.9XXA, X75.XXXA, X76.XXXA, X77.0XXA, X77.1XXA, X77.2XXA, X77.3XXA, X77.8XXA, X77.9XXA, X78.0XXA, X78.1XXA, X78.2XXA, X78.8XXA, X78.9XXA, X79.XXXA, X80.XXXA, X81.0XXA, X81.1XXA, X81.8XXA, X82.0XXA, X82.1XXA, X82.2XXA, X82.8XXA, X83.0XXA, X83.1XXA, X83.2XXA, X83.8XXA |
| **Suicidal ideation** | V62.84 | R45.851 |
| **Suicide attempt** | - | T14.91 |
| **Depression (based on DSM-5)** | | |
| **Disruptive mood dysregulation disorder** | 296.99 | F34.8 |
| **Major depressive disorder, single episode, mild** | 296.21 | F32.0 |
| **Major depressive disorder, single episode, moderate** | 296.22 | F32.1 |
| **Major depressive disorder, single episode, severe** | 296.23 | F32.2 |
| **Major depressive disorder, single episode, with psychotic features** | 296.24 | F32.3 |
| **Major depressive disorder, single episode, in partial remission** | 296.25 | F32.4 |
| **Major depressive disorder, single episode, in full remission** | 296.26 | F32.5 |
| **Major depressive disorder, single episode, unspecified** | 296.2 | F32.9 |
| **Major depressive disorder, recurrent episode, mild** | 296.31 | F33.0 |

| | | |
|---|---|---|
| **Major depressive disorder, recurrent episode, moderate** | 296.32 | F33.1 |
| **Major depressive disorder, recurrent episode, severe** | 296.33 | F33.2 |
| **Major depressive disorder, recurrent episode, with psychotic features** | 296.34 | F33.3 |
| **Major depressive disorder, recurrent episode, in partial remission** | 296.35 | F33.41 |
| **Major depressive disorder, recurrent episode, in full remission** | 296.36 | F33.42 |
| **Major depressive disorder, recurrent episode, unspecified** | 296.3 | F33.9 |
| **Persistent depressive disorder** | 300.4 | F34.1 |
| **Premenstrual dysphoric disorder** | 625.4 | N94.3 |
| **Depressive disorder due to another medical condition** | 293.83 | |
| **With depressive features** | | F06.31 |
| **With major depressive-like episode** | | F06.32 |
| **With mixed features** | | F06.34 |
| **Other specified depressive disorder** | 311 | F32.8 |
| **Unspecified depressive disorder** | 311 | F32.9 |
| **Anxiety disorders (based on DSM-5)** | | |
| **Separation anxiety disorder** | 309.21 | F93.0 |
| **Selective mutism** | 313.23 | F94.0 |
| **Specific phobia** | 300.29 | |
| **Animal** | | F40.218 |
| **Natural environment** | | F40.228 |
| **Fear of blood** | | F40.230 |
| **Fear of injections and transfusions** | | F40.231 |
| **Fear of other medical care** | | F40.232 |
| **Fear of injury** | | F40.233 |
| **Situational** | | F40.248 |

| | | |
|---|---|---|
| **Other** | | F40.298 |
| **Social anxiety disorder (social phobia)** | 300.23 | F40.10 |
| **Panic disorder** | 300.01 | F41.0 |
| **Agoraphobia** | 300.22 | F40.00 |
| **Generalized anxiety disorder** | 300.02 | F41.1 |
| **Anxiety disorder due to another medical condition** | 293.84 | F06.4 |
| **Other specified anxiety disorder** | 300.09 | F41.8 |
| **Unspecified anxiety disorder** | 300 | F41.9 |

**eTable 3: Medication Codes for GLP-1 Receptor Agonists**

| Drug Type | RxNorm | National Drug Codes |
|---|---|---|
| **Albiglutide** | 1534801, 1534763, 1534805, 1534822, 1534802, 1534821, 1659117, 1534804, 1534800, 1534820, 1534797, 1534819, 1659115, 1534798 | 00173086601, 00173086602, 00173086635, 00173086661, 00173086701, 00173086702, 00173086735, 00173086761 |
| **Dulaglutide** | 1551296, 1551291, 1551300, 1551306, 2395779, 2395785, 1551297, 1551305, 2395778, 2395784, 1649586, 1551299, 1551295, 1551304, 2395777, 2395783, 1551292, 1551303, 2395776, 2395782, 1649584, 1551293 | 00002143301, 00002143361, 00002143380, 50090348400, 50090645300, 54568043363, 54568043371, 00002143401, 00002143461, 00002143480, 50090348300, 50090645600, 54568043463, 54568043471, 00002223601, 00002223661, 00002223680, 50090546700, 50090657100, 00002318201, 00002318261, 00002318280 |
| **Exenatide** | 1242964, 604751, 60548, 1242968, 1544918, 1990869, 847913, 847917, 1242965, 1653613, 1990867, 847911, 847916, 1653614, 1653619, 1653625, 1990868, 1169415, 1242967, 1242963, 1544916, 1990866, 847910, 847915, 1242961, 1653610, 1990864, 847908, 847914, 1653611, 1653616, 1990865, 1163790 | 00310652004, 66780021902, 66780021904, 66780022601, 00310653001, 00310653004, 00310653085, 00310654001, 00310654004, 00310654085, 00002021008, 00310652401, 54868538401, 66029021008, 66780021008, 66780021201, 66914103505, 68258894802, 00002021007, 00002021009, 00310651201, 00310651285, 54868538400, 54868538402, 66029021007, 66780021007, 66780021009, 66914103504, 68258894701 |

| | | |
|---|---|---|
| **Liraglutide** | 1598264, 1860168, 897123, 475968, 1727493, 1598268, 1860172, 897126, 1598265, 1860169, 897124, 1653597, 1653600, 1860170, 1186578, 1598267, 1860171, 1860167, 897122, 1860164, 897120, 1653594, 1860166, 1163230, 1860165 | 00169291115, 00169291190, 00169291197, 00169280013, 00169280015, 00169280090, 00169280097, 50090425700, 00169406012, 00169406013, 00169406090, 00169406097, 00169406098, 00169406099, 50090285300, 50090450300, 54569650700 |
| **Lixisenatide** | 1803887, 1858996, 1803903, 1803902, 1440051, 1858994, 1803893, 1803896, 1859000, 1803888, 1803895, 1858997, 1803889, 1858998, 1803890, 1858999, 1803892, 1803894, 1858995, 1440052, 1440056, 1858991, 1803885, 1858993, 1440053, 1858992 | 00024576101, 00024576102, 00024576105, 00024576302, 00024574502, 00024574101, 00024574000, 00024574702, |
| **Semaglutide** | 1991307, 2200645, 2553502, 1991302, 1991311, 1991317, 2200650, 2200654, 2200658, 2398842, 2553506, 2553603, 2553803, 2553903, 2554104, 2599365, 2619154, 1991308, 2200646, 2200653, 2200657, 2553503, 2553602, 2553608, 2553902, 2554103, 2599364, 2619153, 1991309, 2200647, 2553504, 1991310, 2200648, 2200649, 2553505, 1991306, 1991316, 2200644, 2200652, 2200656, 2398841, 2553501, 2553601, 2553802, 2553901, 2554102, 2599362, 2619152, 1991303, 2200640, 2200651, 2200655, 2553400, 2553600, 2553606, 2553900, 2554101, 2599361, 2619151, 1991305, 2200643, 2553500, 1991304, 2200641, 2200642 | 00169413211, 00169413212, 00169413290, 00169413297, 50090513800, 70518214300, 00169413602, 00169413611, 50090513900, 00169431401, 00169431413, 00169431430, 00169430301, 00169430313, 00169430330, 00169430390, 00169430393, 00169430399, 00169430701, 00169430713, 00169430730, 00169413001, 00169413013, 50090594900, 00169452501, 00169452514, 00169452590, 00169452594, 50090582400, 00169450501, 00169450514, 00169450101, 00169450114, 00169451701, 00169451714, 00169452401, 00169452414, 00169477211, 00169477212, 00169477290, 00169477297, 50090605100, 00169418103, 00169418113, 00169418190, 00169418197 |
| **Tirzepatide** | 2601734, 2669702, 2601723, 2601746, 2601758, 2601764, 2601770, 2601776, 2601785, 2644399, 2644403, 2644407, 2644411, 2644415, 2644419, 2669706, 2669709, 2669712, 2669715, 2669718, 2669721, 2601745, 2601757, 2601763, 2601769, 2601775, 2601781, 2669703, 2669708, 2669711, | 00002149501, 00002149580, 00002145701, 00002145780, 00002150601, 00002150661, 00002150680, 00002147101, 00002147180, 00002146001, 00002146080, 00002148401 |

| | |
|---|---|
| | 2669714, 2669717, 2669720, 2601737, 2644398, 2669705, 2601736, 2669704, 2601743, 2601755, 2601761, 2601767, 2601773, 2601784, 2644396, 2644401, 2644405, 2644409, 2644413, 2644417, 2601742, 2601754, 2601760, 2601766, 2601772, 2601778, 2601731, 2644395, 2601730 |
